# Comprehensive Demographic Correction Improves Sensitivity and Reduces Bias in Cognitive Assessment

**DOI:** 10.64898/2026.06.27.26356750

**Authors:** David L. Woods, Kathleen Hall, Isabella Jaramillo, Mike Blank, Kristi Geraci, Peter Pebler, David K. Johnson

## Abstract

**Background:** Scores on neuropsychological assessments are typically corrected for the influences of age, education, and gender (AEG). However, other demographic factors, such as crystallized ability and race/ethnicity, independently affect test performance. As a result, standard scores systematically over- or under-classify impairment in patients whose demographic profile differs from that of the reference population.

**Methods:** We developed a Comprehensive (C-) model scoring algorithm that added vocabulary, age², race/ethnicity, Latino background, a coarse socioeconomic status proxy, computer use, and daily prescription medications to the standard AEG predictor pool. The model was developed using data from 1,914 community-dwelling adults assessed with the California Cognitive Assessment Battery (CCAB; Woods et al., 2024). For each of 118 individual cognitive measures, stability-selection LASSO identified robust predictors in 300 random 80/20 splits retained at ≥ 80% frequency and then estimated mean coefficients and confidence intervals in 1,000 bootstrap OLS samples. Cross-sample frozen-coefficient validation was used to evaluate scoring model generalization in two subgroups: Group 1 (n = 1,033, older, first enrolled cohort) and Group 2 (n = 881, a recently recruited younger cohort).

**Results:** Stability selection retained a mean of 2.81 predictors per measure (range 1-6). Compared to the AEG model, the C-model approximately doubled variance explained (r² = 0.50 vs 0.25; mean across cognitive domains r² = 0.32 vs 0.18) and outperformed AEG in 98.8% of individual measures with non-trivial demographic signal. Racial disparities in MCI classification (the bottom-7th-percentile) were substantially reduced: Black-vs-White ratios fell from 5.6 (AEG) to 1.8 (C). Conversely, sensitivity was improved in individuals with elevated premorbid function: MCI classification ratios in low-vs-high vocabulary quartiles fell from 11.3 to 2.1. AIC favored the C-model in 88.1% of measures (mean ΔAIC = −167), ruling out overfitting. Frozen-coefficient validation preserved the C-model’s r² advantage in every cognitive domain.

**Conclusions:** By correcting scores for race, premorbid cognitive functioning (vocabulary), and other demographic predictors, the C-model explains substantially more variance than the AEG model, reduces racial bias, and increases sensitivity to cognitive decline in high-functioning participants. C and AEG models can be used in parallel: model concordance increases diagnostic confidence, while disagreement carries diagnostic information.

**Highlights:**

- We developed a Comprehensive (C-) model for scoring California Cognitive Assessment Battery (CCAB) tests that supplements standard age + education + gender (AEG) demographic corrections with additional predictors including vocabulary, age², race/ethnicity, Latino background, socioeconomic status (SES), computer use, and daily medications.
- To avoid model overfitting, significant predictors were identified with stability-selection LASSO, resulting in a mean C-model retention of 2.81 (range 1-6) predictors per individual test score.
- In 1,914 community-dwelling adults assessed with CCAB, the C-model approximately doubled explained variance for overall performance (OMNI) scores when compared with the AEG model (r² = 0.50 vs 0.25), and accounted for more variance than the AEG model in 98.8% of measures with non-trivial demographic signal. Cross-sample validation with two demographically distinct cohorts showed that the C-model’s r² advantage was preserved in every cognitive domain (Δr² variation < ±0.020 across fits), supporting generalizability.
- The C-model reduced demographic classification disparities in classifying mild cognitive impairment (MCI, bottom 7% of participants) and normalized MCI detection across different levels of premorbid cognitive reserve: Black-vs-White MCI-classification ratios fell from 5.6 (AEG) to 1.8 (C), and low-vs-high vocabulary performance ratios fell from 11.3 to 2.1.
- C-model scores are orthogonal to vocabulary; comparisons with premorbid crystallized intelligence are incorporated directly in the scoring algorithm, eliminating the need for post-hoc comparisons.
- The C-model is best used in parallel with AEG scoring: concordance between models increases diagnostic confidence, while disagreement provides additional diagnostic information.

## 1. Introduction

### 1.1 Demographic correction of cognitive test scores

Cognitive tests are scored relative to expected results from neurologically normal control participants with similar demographic backgrounds based on demographic score correction: raw scores are converted to standard scores adjusted for demographic factors predictive of performance. The dominant convention—corrections for age, education, and gender (AEG)—was established by the Heaton norms (Heaton, Grant, & Matthews, 1991) and persists in all major batteries: the Wechsler scales, the Halstead-Reitan tradition, the NIH Toolbox, and the NACC Uniform Data Set. AEG correction is typically implemented as a continuous regression-based adjustment with predicted scores generated from population means to characterize a patient’s standing relative to demographically matched peers.

AEG correction is computationally simple, clinically familiar, and adequate for many assessment purposes. But it is also incomplete in ways that have become increasingly difficult to ignore as cognitive testing has expanded to more demographically heterogeneous populations and as computational statistical methods enabling richer normative modeling have matured.

### 1.2 Why AEG is incomplete

Two convergent critiques question whether AEG correction remains adequate as the field’s default.

#### Demographic distortion

Cognitive test performance correlates with race, vocabulary, socioeconomic status, computer familiarity, and medical comorbidity in ways that AEG does not capture (Manly, Jacobs, Touradji, Small, & Stern, 2002; Manly & Echemendia, 2007). As a result, AEG-corrected scores retain demographic structure that distorts clinical classification, particularly in patients whose demographic profile differs from the normative reference population (for example, biasing AEG scores for Black, Asian, and Hispanic participants underrepresented in most normative AEG datasets). In addition, low-vocabulary patients may be over-classified as cognitively impaired by AEG cutoffs at rates that do not reflect the population prevalence of pathology.

#### The crystallized–fluid tradition

Clinical neuropsychology has long used vocabulary or word-reading as a proxy of premorbid ability against which current performance is compared (Nelson, 1982; Grober & Sliwinski, 1991; Wechsler, 2001, 2009). The National Adult Reading Test (NART), American National Adult Reading Test (AMNART), Wechsler Test of Adult Reading (WTAR), and Test of Premorbid Functioning (TOPF) all operationalize this approach, predicting premorbid IQ from word-reading performance and thus enabling the clinician to compare the scores from the cognitive assessment with scores predicted based on premorbid IQ. This crystallized–fluid comparison is central to the clinical assessment of age-related cognitive decline, but requires an additional correction: estimating how a normal subject, with similar demographic background and premorbid IQ, would have performed on the test. Typically, test norms and premorbid IQ corrections derive from different population samples, limiting the precision and validity of the comparison.

### 1.3 Comprehensive demographic correction

Computational advances have made comprehensive demographic correction viable for two reasons. First, statistical methods now exist that allow principled extension of AEG to a richer predictor set without overfitting. LASSO regression with stability selection (Meinshausen & Bühlmann, 2010) identifies only predictors which reliably influence a score, using strict selection thresholds to avoid overfitting. Bootstrap estimation of the coefficients of the surviving predictors can then provide robust confidence intervals for each coefficient. Second, computerized testing enables automated or semi-automated collection of test performance, including demographic data and premorbid ability, and produces large, well-characterized normative samples that support advanced modeling. These approaches enable scoring pipelines to include richer demographic correction and integrate crystallized-fluid comparisons.

### 1.4 Aims and approach

This paper has four aims:

#### Aim 1

Develop a Comprehensive (C-) model of cognitive normative scoring that adds vocabulary (as premorbid ability proxy), age² (residualized), race/ethnicity, Latino background, socioeconomic status (SES), computer use, and daily medication use to the standard AEG predictor pool, then use stability-selection LASSO to retain only significant predictors, followed by coefficients bootstrap estimation to provide robust coefficient estimates and confidence intervals for each score.

#### Aim 2

Compare C-model scores with AEG-model scores to evaluate (a) reduction of demographic classification biases, (b) variance explained, and (c) model parsimony.

#### Aim 3

Test AEG- and C-model generalization using held-out cross-sample validation in two demographically distinct subsample populations (older / younger) by comparing AEG- and C-models’ out-of-sample performance.

#### Aim 4

Provide a clinical decision framework for integrating AEG-corrected, and C-model corrected scores to optimize diagnosis and interpret ambiguous cases.

## 2. Methods

### 2.1 Participants

The normative sample comprised 1,914 community-dwelling adults assessed in the San Francisco Bay Area from 2021 to 2026. Demographic characteristics are shown in Table 1. Mean age was 53.1 years (SD 17.3, range 18–94); 56.6% were female; the racial composition was 38.1% White, 22.6% Black, 17.9% Asian, and 22.3% Other or Mixed race; 15.6% identified as Hispanic/Latino. All participants provided written informed consent under protocols approved by the Western Institutional Review Board.

**Table 1.** Demographic characteristics of the full normative sample and the two recruitment cohorts. Test statistics compare Group 1 to Group 2. Race categories (White, Black, Asian, Other/Mixed) were mutually exclusive, with Hispanic/Latino coded separately. Group comparisons are Welch’s t-tests for continuous variables and Pearson χ² (uncorrected) for proportions.

| Characteristic | Full sample (n = 1,914) | Group 1 (n = 1,033) | Group 2 (n = 881) | G1 vs G2 test |
| --- | --- | --- | --- | --- |
| Age, years, M (SD) | 53.11 (17.29) | 62.20 (13.77) | 42.46 (14.74) | t = 30.11, p < .001 |
| Age range, years | 18–89 | 18–89 | 18–87 | — |
| Female, % | 56.6 | 56.2 | 57.1 | $\chi^2 = 0.14$, p = .708, ns |
| Education, years, M (SD) | 15.54 (2.29) | 15.84 (2.36) | 15.18 (2.16) | t = 6.26, p < .001 |
| White, % | 38.1 | 38.5 | 37.6 | $\chi^2 = 0.18$, p = .667, ns |
| Black, % | 22.6 | 27.7 | 16.6 | $\chi^2 = 33.61$, p < .001 |
| Asian, % | 17.9 | 18.4 | 17.4 | $\chi^2 = 0.34$, p = .560, ns |
| Other / Mixed race, % | 22.3 | 17.0 | 28.5 | $\chi^2 = 35.98$, p < .001 |
| Hispanic / Latino, % | 15.6 | 13.0 | 18.6 | $\chi^2 = 11.52$, p < .001 |
| Vocabulary score, M (SD) | 34.56 (9.36) | 35.80 (9.45) | 33.10 (9.05) | t = 6.38, p < .001 |
| Daily medications, M (SD) | 1.61 (2.00) | 2.09 (2.08) | 1.04 (1.75) | t = 11.97, p < .001 |

Note. ns = not significant at p < .05. Vocabulary score is the CCAB Vocabulary subtest raw score.

The sample comprised two recruitment cohorts that were independently shown to fit the same confirmatory bifactor factor structure across cognitive parcels (Woods et al., companion manuscript, in preparation). Group 1 (n = 1,033) an older cohort of early recruits that completed three days of testing at enrollment (mean age 62.2, SD 13.8). Group 2 (n = 881) was a more recently recruited younger cohort (mean age 42.5, SD 14.7) with a higher proportion of participants who self-identified as Other/Mixed race (28.5% vs 17.0%) and of Hispanic/Latino background (18.6% vs 13.0%).

### 2.2 Cognitive assessment: the California Cognitive Assessment Battery (CCAB)

Cognitive performance was assessed with the California Cognitive Assessment Battery (CCAB; Woods et al., 2024), an examiner-proctored, self-administered computerized battery that requires approximately 130 minutes to complete. Assessments were telemedically proctored and administered in participants’ homes. The test battery included a brief (4-minute) vocabulary test, which was used as a proxy for crystallized intelligence and a covariate in the Comprehensive model, and a questionnaire that obtained detailed demographic information. Participants completed 24 CCAB subtests that generated 118 computer-extracted measures, including speech and language biomarkers (SLBs). Scores were grouped into five cognitive domains: Executive Function (EF), Memory (MEM), Language/Semantic (LS), Processing Speed (PS), and Speech Fluency (SF), plus an omnibus composite score reflecting overall cognitive performance on all tests.

### 2.3 Score derivation pipeline

Scores were automatically extracted from all tests after speech transcription with Consensus Automatic Speech Recognition (CASR; Woods et al., 2025). . Raw scores were processed through an identical pipeline before normative correction: (1) winsorization at ±3 SD; (2) imputation of remaining missing values; (3) log-transformation and inversion of temporal measures (e.g., response time, decision time, inter-word intervals, etc.) so that higher z-scores consistently indicated better performance; (4) z-scoring against the full normative sample; (5) a second winsorization on z-scores at ±3.5 SD to handle outliers in transformed measures; and (6) parcel-level averaging to produce domain composites and the OMNI (overall) composite.

### 2.4 AEG model scoring

For each measure, we fitted bootstrap ordinary least squares (OLS) regressions of the form score ∼ age + education + gender (female), with 1,000 bootstrap samples to obtain mean coefficients and 90% percentile confidence intervals. The mean coefficients were then applied to the full sample to produce predicted values; AEG-residual scores were obtained as observed minus predicted. This implementation replicates the conventional AEG approach used in the Heaton norms, NIH Toolbox, and NACC UDS.

### 2.5 C-model scoring

#### Predictor pool

The C-model considers eight candidate predictors: age, age²-residual (to quantify non-linear age effects), education, gender, race (Black, Asian, Other; reference category = White), Latino background, vocabulary, a coarse SES proxy (constructed from marital status and number of rooms in residence), daily computer use (to estimate intellectual engagement), and daily medication count (to quantify comorbidities).

#### Stability selection

For each measure, predictor selection used LASSO regression at a strict (lambda.1se) penalty threshold across 300 random 80/20 sample splits. A predictor was retained only if (a) it survived the lambda.1se threshold in at least 80% of the 300 splits (Meinshausen & Bühlmann, 2010), and (ib) its bootstrap-mean standardized coefficient reached |β| ≥ 0.10 — a magnitude threshold ensuring that retained predictors had meaningful effects. Race indicators were handled with an all-or-none rule: if any of the three race coefficients (Black, Asian, Other) passed selection for a given measure, all three were retained. This avoids selective per-group adjustments that would be difficult to interpret.

#### Coefficient estimation and scoring

Once the predictor set was stability-selected for a given measure, mean coefficients were estimated from 1,000 bootstrap OLS samples fitted to the selected formula. For each retained predictor, the mean β weights and 95% confidence intervals (mean ± 1.96 SD across bootstrap samples) were reported. Mean coefficients were applied to the full sample to generate predicted values, and C-model-residualized scores were computed as observed minus predicted.

### 2.6 Validation

Cross-sample validation used a frozen-coefficient design. The C-model was fit on Group 1 only (n = 1,033) using the full LASSO + stability selection + bootstrap pipeline; the resulting mean coefficients were then applied to all 1,914 subjects to produce predictions and residuals. The C-model was then fit on Group 2 only (n = 881) and its frozen coefficients applied to all 1,914 subjects. The same fit-and-apply procedure was performed on the AEG model. This design produces in-sample predictions for subjects in the fit subgroup and held-out predictions for subjects not in the fit subgroup, allowing the calculation of validation r² and predictor-stability statistics across cohorts.

### 2.7 Statistical analyses

Primary analyses comprised:

#### Fit statistics

We compared AEG vs C-model scores for adjusted r², RMSE, Akaike Information Criterion (AIC) for model parsimony, and Kolmogorov-Smirnov, Anderson-Darling, and Shapiro-Wilk tests of residual normality. Predictor retention rates and coefficient stability were analyzed across the full-sample, fit-on-G1, and fit-on-G2 runs. Held-out validation r² shrinkage, computed as the difference between in-sample r² (fit subjects) and held-out r² (non-fit subjects) was evaluated for each fit.

MCI classification rates were computed for both correction methods (AEG, C-model) using the within-distribution 7th percentile cutoff to approximate the z = −1.5 cutoff used clinically to flag MCI-range performance. We used the within-distribution percentile rather than a fixed z-cutoff to control for the reduced residual variance of omnibus and cognitive domain scores.

## 3. Results

### 3.1 Sample characteristics

Demographic characteristics of the full sample, Group 1, and Group 2 are presented in Table 1. Group 1 was substantially older than Group 2 (mean ages 62.2 vs 42.5, t = 30.11, p < .001), more often Black (27.7% vs 16.6%, χ² = 32.98, p < .001), more medicated (mean 2.09 vs 1.04 daily medications, t = 11.97, p < .001), and had larger vocabularies (35.8 vs 33.1, t = 6.38, p < .001), consistent with the well-documented increase of vocabulary across the adult lifespan (Verhaeghen, 2003; Kavé, 2024). Group 2 was more often Hispanic/Latino (18.6% vs 13.0%) and more often Other/Mixed race (28.5% vs 17.0%). Education years (15.84 vs 15.54), Gender (56.2% vs 57.1% female) and Asian race (18.4% vs 17.4%) did not differ between groups. The substantial age difference between Groups 1 and 2 makes the cross-sample validation a more demanding test of C-model generalizability than within-sample resampling would provide.

### 3.2 Demographic bias

Figures 1a shows mean cognitive z-scores (omnibus and cognitive domain score for White and non-White race under each correction method. Both uncorrected and AEG-corrected scores produced strong demographic gradients: White > Non-White on every domain, with the largest absolute separation (∼0.5 SD) seen when using the AEG model. Figures 1b shows mean cognitive z-scores by vocabulary quintile. Individuals in the top quintile outperformed those in the bottom quintile by more than 1.0 SD for the AEG mode. The C-model eliminated both race and vocabulary gradients, producing nearly identical scores across race and vocabulary groups in all cognitive domains.

**Figure 1a.**
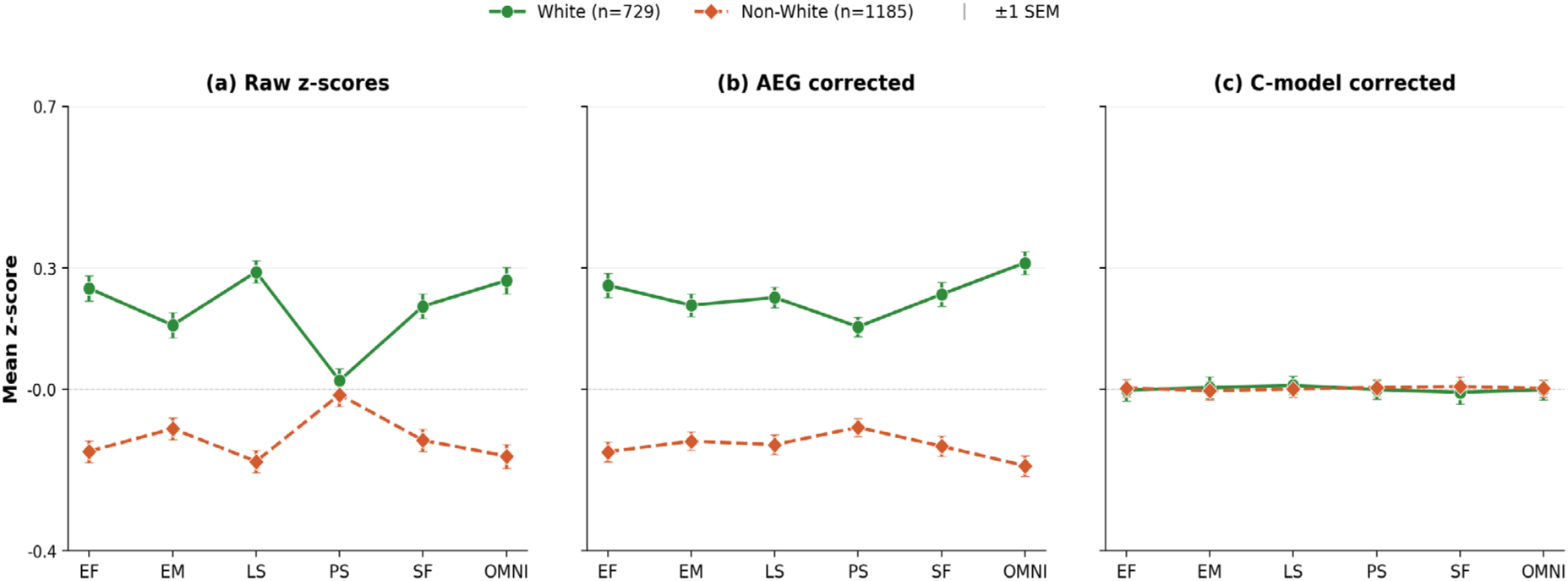
Mean cognitive z-scores for White vs Non-White group membership across five cognitive domains and the omnibus composite: (a) Raw (uncorrected) z-scores show a White > Non-White gap of approximately 0.3–0.5 SD across most domains. (b) AEG-corrected scores leave similar gaps in most domains (correcting for age increases the racial gap in Processing Speed because the non-White group was younger). (c) C-model scores corrected for race eliminating the gaps on every domain. Error bars = ±1 SEM. EF = Executive Function; EM = Episodic Memory; LS = Language/Lexical; PS = Processing Speed; SF = Semantic Fluency; OMNI = omnibus composite (mean all tests).

**Figure 1b.**
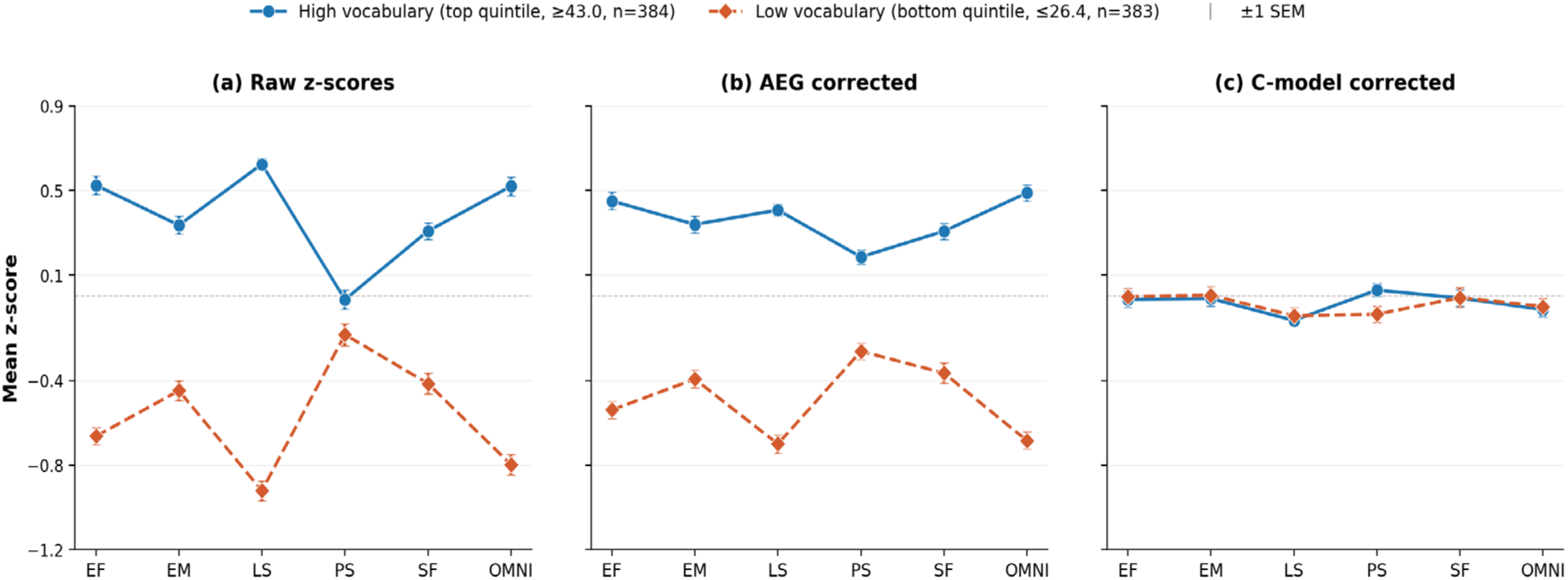
Mean cognitive z-scores comparing performance of participants with vocabularies in the top vs bottom quintiles across five cognitive domains and the omnibus composite. (a) Uncorrected z-scores show a high-vs-low vocabulary gap exceeding 1.0 SD in several domains. (b) AEG-corrected scores slightly reduced most gap but large vocabulary-related differences remained (∼0.9–1.0 SD for the omnibus score). (c) C-model scores eliminated the gaps, producing residuals that were orthogonal to vocabulary by construction. Error bars = ±1 SEM.

Table 2 provides detailed MCI clinical classification rates (the bottom-7th-percentile) for omnibus scores using the different scoring models. Uncorrected z-scores classified 16.0% of Black subjects in the MCI group versus 3.1% of White subjects. AEG correction slightly increased this disparity, resulting in a 5.6-fold ratio of MCI prevalence in Black (14.8%) vs. White (2.7%) participants. The C-model reduced the Black/White ratio to 1.8 (9.0% vs 5.1%), removing the bulk of the demographically-driven disparity while preserving a smaller residual difference that likely reflected cognitive risk-factor exposure differences.

**Table 2.**
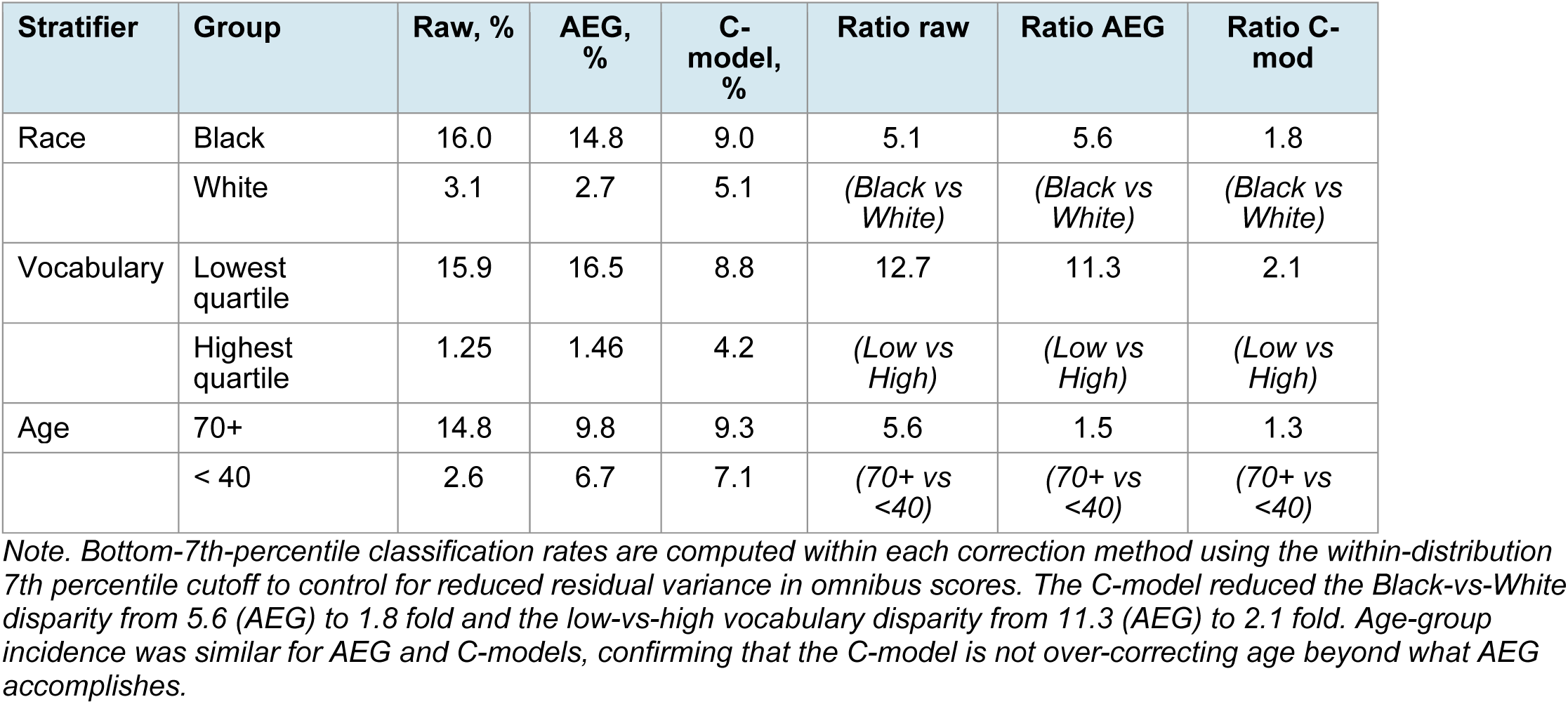
Bottom-7th-percentile (MCI-range) classification rates on the OMNI composite by race, vocabulary quartile, and age group, under three correction methods. Disparity ratios are computed within method (e.g., 16.0% / 3.1% = 5.1× for the raw Black-vs-White comparison). Reductions in disparity ratios between AEG and the C-model are reported in the rightmost column.

| Stratifier | Group | Raw, % | AEG, % | C-model, % | Ratio raw | Ratio AEG | Ratio C-mod |
| --- | --- | --- | --- | --- | --- | --- | --- |
| Race | Black | 16.0 | 14.8 | 9.0 | 5.1 | 5.6 | 1.8 |
|  | White | 3.1 | 2.7 | 5.1 | (Black vs White) | (Black vs White) | (Black vs White) |
| Vocabulary | Lowest quartile | 15.9 | 16.5 | 8.8 | 12.7 | 11.3 | 2.1 |
|  | Highest quartile | 1.25 | 1.46 | 4.2 | (Low vs High) | (Low vs High) | (Low vs High) |
| Age | 70+ | 14.8 | 9.8 | 9.3 | 5.6 | 1.5 | 1.3 |
|  | < 40 | 2.6 | 6.7 | 7.1 | (70+ vs <40) | (70+ vs <40) | (70+ vs <40) |

*Note.* Bottom-7th-percentile classification rates are computed within each correction method using the within-distribution 7th percentile cutoff to control for reduced residual variance in omnibus scores. The C-model reduced the Black-vs-White disparity from 5.6 (AEG) to 1.8 fold and the low-vs-high vocabulary disparity from 11.3 (AEG) to 2.1 fold. Age-group incidence was similar for AEG and C-models, confirming that the C-model is not over-correcting age beyond what AEG accomplishes.

Score disparities reflecting vocabulary estimates of premorbid IQ were even more dramatic. Among subjects in the lowest vocabulary quartile, AEG-based z-scores classified 16.5% as MCI and among the highest quartile, only 1.46%. This 11-fold disparity suggests that AEG scores were insensitive to MCI in individuals with high levels of cognitive reserve. The C-model reduced the disparity to 2.1-fold (8.8% vs 4.2%), a base-rate more consistent with realistic population-level MCI prevalence estimates, reflecting the crystallized–fluid correction: a high-vocabulary individuals with fluid performance well below their premorbid baseline were flagged by the C-model even when their AEG corrected fluid performance fell within the population mean range.

### 3.3 Crystallized–fluid orthogonality

The C-model contains the crystallized-fluid correction framework in the scoring algorithm itself, correcting for vocabulary on scores where vocabulary significantly influenced performance. Thus, by construction, C-model corrected scores are orthogonal to vocabulary. Table 3 reports Pearson correlations of domain-corrected scores with vocabulary scores across the three correction methods.

**Table 3.** Pearson correlations of cognitive scores with vocabulary across the OMNI composite and five cognitive domains, under three correction methods (n = 1,914).

| Domain | Raw r | AEG r | C-model r | Domain type |
| --- | --- | --- | --- | --- |
| OMNI | +0.47 | +0.49 | **+0.001** | *composite* |
| LS | +0.64 | +0.50 | **0.000** | *crystallized* |
| EF | +0.46 | +0.42 | **+0.002** | *fluid* |
| EM | +0.31 | +0.33 | **0.000** | *fluid* |
| SF | +0.28 | +0.27 | **0.000** | *mixed* |
| PS | +0.05 | +0.21 | **+0.06** | *fluid* |

*Note.* AEG-corrected scores retain substantial correlations with vocabulary ( $r = 0.21$ – $0.50$ across domains) despite correction for education. C-model correction reduces vocabulary correlations to essentially zero. AEG correction increases vocabulary's correlation with processing speed that is suppressed by age in raw PS scores; once AEG removes age, the vocabulary correlation is exposed. The C-model adjusts for both, restoring near-orthogonality.

Age-group disparities followed a similar pattern. Raw z-scores classified 14.8% of subjects aged 70+ in the bottom 7% versus 2.6% of subjects under 40. AEG correction substantially reduced the bias (9.8% vs 6.7%, 1.5-fold) because age was a predictor. The C-model further regularized the age-group ratio (9.3% vs 7.1%, 1.3-fold).

Three findings warrant attention. First, raw cognitive scores correlated substantially with vocabulary across all five domains (r = +0.28 to +0.64) and the omnibus composite (r = +0.47), confirming the well-established crystallized-fluid covariance in the general population. Second, AEG correction left these correlations almost entirely intact (r = +0.27 to +0.50; OMNI r = +0.49): vocabulary-cognition covariance was largely independent of education and hence is preserved after AEG regression. Third, by construction, C-model correction reduced the vocabulary correlation to essentially zero across all domains.

The processing speed (PS) result is particularly informative as a methodological caveat. Raw PS correlated with vocabulary at only r = +0.05—not because vocabulary is unrelated to PS, but because the strong age effect on PS (raw r ≈ −0.61 with age) suppressed the underlying vocabulary correlation. Because Vocabulary was positively corrected with Age (r = 0.22), by removing the age effect, AEG correction exposed a spurious correlation with Vocabulary (r = +0.21) in PS scores. The C-model, by correcting for both age and vocabulary, restored near-orthogonality (r = +0.06).

### 3.4 Variance explained

Figure 3 presents the comparison of variance explained and residual error for AEG and C-model fits for omnibus and domain composite scores. Compared to the AEG model, the C-model approximately doubled the AEG r² for the OMNI composite (0.50 vs 0.25, Δr² = +0.25). RMSE fell correspondingly (0.71 vs 0.87, Δ = −0.16), implying that 95% prediction intervals on patient-level corrected scores were approximately 18% narrower with the C-model vs. the AEG model.

**Figure 3.**
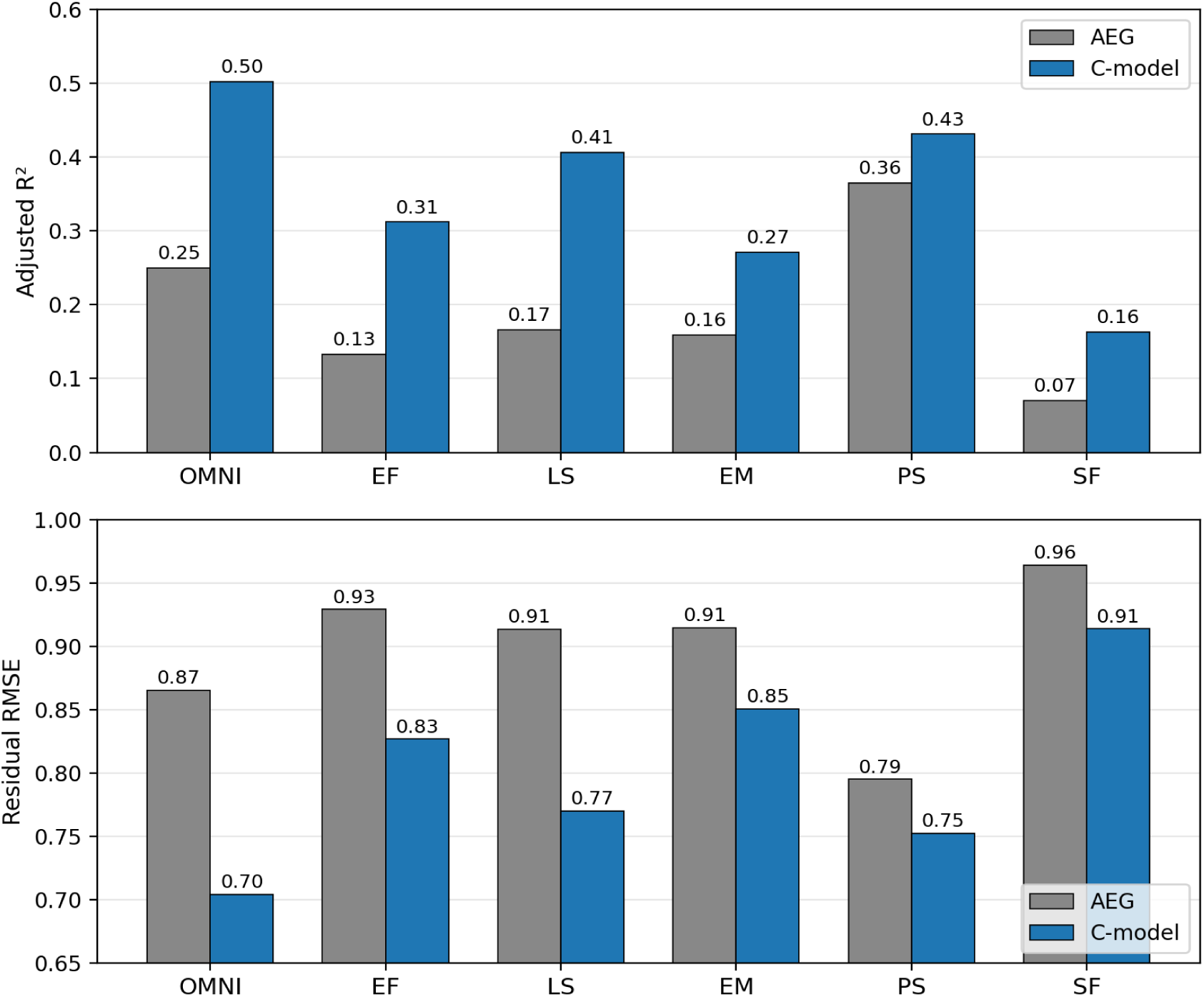
Comparison of AEG and C-model demographic correction across the OMNI composite and five cognitive domains (n = 1,914; bootstrapped with 300 iterations). Top panel: adjusted r² (higher = better fit). Bottom panel: residual RMSE (lower = better fit). The C-model approximately doubles AEG r² for omnibus (OMN) scores (0.50 vs 0.25, Δ = +0.25) and produces substantial gains in every domain, with the largest absolute improvements in domains most heavily loaded on cultural and educational exposure factors (LS, EF). RMSE falls correspondingly producing narrower patient-level prediction intervals with the C-model.

Domain-level improvements of the C-model fit were also substantial and followed a theoretically interpretable pattern. The largest absolute r² improvements occurred in language/lexical (LS: +0.24) and executive function domains (EF: +0.18) that are most heavily loaded on premorbid cognitive ability that the AEG model does not capture. Episodic memory (EM: +0.11) and Speech fluency (SF: +0.09) showed moderate gains. The smallest gain was in processing speed (PS: +0.07), where AEG already captured the dominant predictor, age (AEG r² = 0.37, the best AEG fit among the five domains) leaving the C-model with less range for improvement.

PS, is also methodologically informative for another reason. Although AEG correction of PS achieved r² = 0.37, well above its fit in any other domain, the C-model still added r² of +0.07 above this strong baseline, with the additional variance explained primarily through vocabulary, daily medications, and computer use, demonstrating that the C-model adds variance proportional to the demographic structure that AEG misses.

#### 3.4.1 Individual test measures

For comparisons restricted to the 70.3% of individual measures with significant demographic signal (AEG adjusted r² > 0.05), the C-model outperformed AEG in 82 of 83 measures (98.8%) by adjusted r². For these measures, the C-model explained roughly 1.6 times as much variance (mean adjusted R² = 0.23 vs 0.14). In other words, in measures where demographic factors meaningfully predicted performance, the C-model was nearly always superior.

### 3.5 C-model parsimony

Stability-selection LASSO produced parsimonious models. Across the 118 measures, the mean number of retained predictors (counting Race as a single factor) was 2.81 (range 1–6) — marginally less than AEG’s mandatory three. Three predictors dominated retention: vocabulary (retained in 72.9% of measures), age (66.9%), and race (61.0%). Gender was retained in 31.4% of measures while education was retained in only 11.9%. The remaining six candidates—Latino background, daily medications, age², SES, computer use—were retained in fewer than 15% of measures each. Socioeconomic status (SES) was a notable case: LASSO at the lambda.1se penalty selected SES with non-zero coefficients in 12 measures (most consistently for Design Fluency, Pause Recall, and Trail Making B). However, its mean coefficient in 1,000 random bootstrap samples never reached |β| ≥ 0.10. SES therefore carried real independent signal after accounting for vocabulary, age, and other demographics, but its magnitude was too small for inclusion in the scoring equation.

Of particular note is the low (11.9%) retention rate for education. With vocabulary as a covariate, education dropped out as a significant predictor in nearly 90% of measures because vocabulary supplanted education as a premorbid ability index. This is consistent with the literacy-vs-education-years literature (Manly et al., 2002; Dotson, Kitner-Triolo, Evans, & Zonderman, 2009; Kotsis, Bobrowicz, & Lambon Ralph, 2025): years of schooling is a poorer marker of cognitive resource than more direct measures of crystallized ability.

#### 3.5.1 Model overfitting

The C-model used LASSO selection to ensure that predictors influenced scores, imposing age as a fallback predictor in 4.1% of scores where LASSO failed to identify any significant predictors. Excluding those cases, C-model predictors were always statistically justified, with confidence intervals never bounding 0.0. In contrast, the AEG forced the fitting of three predictors (age, education, and gender) regardless of whether the predictors carried significant information. Examination of AEG coefficients across the 1,000 bootstrap samples revealed that 49.2% of equations contained at least one coefficient whose 90% confidence interval included 0.0; i.e., could not be distinguished from sampling noise. Insignificant coefficients were most commonly gender (34.7%), but were also seen for age (16.1%) and education (7.6%). Thus, while the AEG model always included three predictors by design, overfitting was common: predictors often lacked statistical support for their inclusion.

Across the full set of 118 measures, AIC favored the C-model in 88.1% of measures with a mean ΔAIC of −167 (median ΔAIC = −113). The AIC advantage is particularly informative because AIC explicitly penalizes additional model parameters: a larger model is favored by AIC only if its variance-explained gain exceeds twice the additional parameter count. The C-model’s AIC superiority therefore rules out the standard concern that ‘more predictors always produce better fit’ as an explanation for its r² advantage.

### 3.6 Model normality

Residual normality was modestly improved under the C-model. Mean Anderson-Darling A statistics were 13.26 for C-model residuals versus 14.97 for AEG residuals (lower = more normal); mean Shapiro-Wilk W was 0.967 vs 0.965; mean Kolmogorov-Smirnov D was 0.055 vs 0.057. The fraction of measures whose residuals passed Shapiro-Wilk at α = .05 was 4.2% under C-model versus 1.7% under AEG—both small in absolute terms (most cognitive measures have non-normal residual distributions due to skew, ceiling effects, and reaction-time distributions), but with the C-model achieving normality at 2.5 times the rate of AEG. The C-model’s removal of additional demographic structure left residuals that were correspondingly cleaner.

### 3.7 Cross-sample validation

Cross-sample validation was performed for AEG and C-model fits on Group 1 (n = 1,033) and Group 2 (n = 881), with frozen coefficients applied to all 1,914 subjects in each fit. Both methods generalized well across cohorts: pooled r² values fell within 0.03 of the full-sample across all five cognitive domains and the omnibus composite for both methods. For the omnibus measure, the AEG full-sample r² was 0.250, Group 1 r² was 0.248, and Group 2 r² applied to all subjects was 0.248. The C-model explained roughly twice the variance in each comparison: the C-model omnibus full-sample r² was 0.501, Group 1 r² was 0.504, and Group 2 r² was 0.500.

The C-model’s r² advantage over AEG was equally evident out-of-sample. The omnibus score advantage was +0.251 in the full-sample fit, +0.256 for the model fit on Group 1 only, and +0.252 for the model fit on Group 2 only. The same pattern held for every cognitive domain: Δr² (C minus AEG) demonstrated that the C-model explained up to 2.5 times the variance of the AEG model, and varied by less than ±0.020 across full, Group 1 fit, and Group 2 fits The C-model’s fit advantage was therefore not an artifact of in-sample optimism that disappeared under held-out testing; it reflected genuine demographic structure that generalized cross-sample.

C-model predictor retention rates were generally stable across fits. Across the three runs (full, fitG1, fitG2), vocabulary was retained in 73%, 74%, and 62% of measures; age in 67%, 65%, 70%; the race indicator set in 61%, 60%, 51%. For predictors retained in both subsamples, mean coefficient values correlated highly across fits: r = +0.77 for vocabulary (n = 67 measures retained in both fits), +0.89 for age (n = 65), and +0.93 for gender (n = 21).

AEG coefficient stability was high but uneven. Age and education coefficients correlated strongly across fits (r = 0.92 and 0.89 respectively), while the gender coefficient was less stable (r = 0.70), and the intercept was sample-dependent (r = −0.229), as expected given the substantial mean-age difference between Group 1 and Group 2.

### 3.8 Score-level generalization

Analysis revealed that C-model predictor-selection showed minor inconsistencies across sub-cohorts, with minimal impact on scores. Comparisons of full C-model scores with scores from Group 1 and Group 2 showed that the median per-subject residual-score correlation between the full model and Group 1 and Group 2 models was r = 0.991 and r = 0.987, with mean per-subject score differences of 0.15 SD and 0.18 SD, and omnibus score differences of 0.11 SD and 0.14 SD. Each of the three models independently flagged 135 of 1,914 subjects at the bottom-7th-percentile OMNI cutoff. The classification disagreement against the full C-model was 1.8% (Group 1) and 1.4% (Group 2), reflecting two borderline cases sitting at a median of 0.05 SD from the cutoff.

The same subject-level generalizability analysis showed that AEG scores were marginally more generalizable across the two subsamples than C-model scores: with median subject-level residual-score correlation between the Group 1 and Group 2 model fits of r = 0.993 for AEG versus r = 0.972 for the C-models. Because the AEG model applied a fixed three-predictor specification without selection, only coefficient magnitudes could differ, leaving less room for cross-group divergence. In contrast, the C-model objectively selected predictors for each measure; as a result both predictors and weights could potentially vary, reducing generalization. Thus, the C-model sacrificed some cross-population consistency (a correlation of 0.97 vs. 0.99) in return for approximately doubling explained variance (omnibus adjusted r² ≈ 0.50 versus 0.25) and reducing demographic disparities in classification (Section 3.4).

These results reconcile the apparent inconsistency between the occasional C-model selection of different predictor sets for Group 1 and Group 2. Because candidate predictors are correlated, alternate fits occasionally selected different sets of related predictors. For example, in a younger group, both education and vocabulary may be retained, while in an older group, only vocabulary. In such cases, the predicted score barely moves when models from different groups were used, because the predictors (vocabulary and education) themselves are significantly correlated, adjusting coefficient magnitudes. Therefore, the corrected scores and abnormality classification remained stable, even where the scoring equations contain different numbers of predictors. The largest scoring model discrepancies among subsamples were concentrated in the language measures where vocabulary, education, and race showed the greatest tradeoffs; even there, subject-level score correlations between independent models remained at or above 0.95.

### 3.9 MCI-level performance

Across the full normative sample, AEG and C-model scores were highly correlated (Pearson r = 0.82 for omnibus; range 0.82–0.94 across the five cognitive domains), but the correlation dropped sharply (r = 0.43 for omnibus; range 0.28–0.67 across the five domains) among participants at-risk of MCI (bottom-7th-percentile z-score). As a result, disagreement was most evident where clinical classification is most important.

Among the all participants who showed MCI-level performance (bottom 7%) on either the C-model or standard model:

- 47.5% (mean age 55.6) showed abnormalities in both C-model and AEG-model omnibus scores (mean AEG score = -2.24; mean C-model score = -2.28).
- 26.2% (mean age 51.4) had abnormal omnibus AEG-model scores but C-model scores in the low normal range (mean AEG score = -1.81; mean C-model score = -0.94). These participants had small vocabularies (83% in the lowest quartile) and were disproportionately Black (50% vs. 22.6% of the cohort).
- 26.2% (mean age 50.5) had omnibus AEG-model scores in the low normal range but abnormal C-model scores (standard score = -1.07; C-model score = -1.76). These participants had large vocabularies (only 4% in the lowest vocabulary quartile) and were disproportionately White (52.1% vs. 38.1% of the cohort).

## 4. Discussion

The continued dominance of age–education–gender (AEG) correction in clinical neuropsychology reflects practical considerations rather than confidence that these three variables provide optimal normative scores. Most widely used neuropsychological batteries were developed during the 1980s and 1990s, when normative scoring systems were designed for printed manuals or simple software implementations. Regression equations with three predictors were computationally straightforward, easily documented, and readily implemented in clinical practice. Consequently, age, education, and gender became the de facto standard across major assessment batteries and have remained so largely through continuity of clinical practice.

Several developments now make more comprehensive demographic correction feasible. First, computerized cognitive assessment batteries can capture demographic variables, medical history, and measures of premorbid ability without increasing administration time. Second, large normative databases provide sufficient statistical power to accurately estimate more complex regression models. Third, modern regularization methods such as stability-selection LASSO permit the objective identification of meaningful predictors from larger sets of potentially relevant factors, while avoiding the inclusion of insignificant predictors. Finally, computerized testing/scoring platforms can automatically apply regression equations, eliminating the practical constraints that once favored simple models. Together, these advances allow normative scoring to move beyond historically constrained AEG models with data-driven demographic corrections that more fully account for the determinants of normal cognitive performance.

### 4.1 Principal findings

The C-model substantially improved on AEG-model scoring in three ways: (1) it approximately doubled the variance explained while improving AIC and residual normality; (2) it reduced demographic classification disparities by 70–85% relative to raw and AEG scores; (3) it produced residuals that were orthogonal to vocabulary. These improvements were achieved with a model that was more parsimonious (mean 2.81 LASSO-selected predictors per measure) than the 3-factor AEG model.

### 4.2 Integrating the crystallized–fluid comparison in the scoring pipeline

The C-model integrated the crystallized–fluid premorbid comparison directly into the scoring pipeline, using co-normed estimates of the influence of vocabulary on test scores. Ideally, a clinician administering a battery such as the WAIS-IV or NIH Toolbox would compare corrected scores against an independent measure of premorbid ability (e.g., the NART or WTAR, Dykiert et al., 2013), but this comparison is often skipped. As a result, a patient with a high premorbid baseline who has domain or omnibus z-scores of -1.0, may often be overlooked, and a patient with a low premorbid baseline and domain or omnibus z-scores of -1.5 may be over diagnosed with MCI.

The C-model integrates the crystallized-vs-fluid comparison directly into the scoring pipeline using co-normed vocabulary scores. Vocabulary is central to the cognitive-reserve framework (Stern, 2009, 2012) and has two properties that make it suitable as a covariate estimate of crystallized abilities. First, it empirically dissociates from years of education (Boyle et al., 2021). In C-model fits, the 11.9% retention rate of education versus the 72.9% retention of vocabulary, reflects this dissociation. Second, vocabulary increases with age across the adult lifespan (Verhaeghen, 2003; Kavé, 2024), opposite to the trajectory of the fluid abilities most affected by acquired cognitive impairment (Salthouse, 2019; Hartshorne & Germine, 2015), making vocabulary suitable as a premorbid baseline marker in older patients whose fluid abilities have declined.

### 4.3 Implications for clinical interpretation

The scoring pipeline of the CCAB reports AEG- and C-model scores for every test because the pattern of agreement and disagreement adds diagnostic information and confidence. For example, when AEG and C-model classifications agree—both flag abnormal performance, or neither does—diagnostic confidence is increased. When performance lands in MCI range for AEG-model scores but not C-model scores, the most likely explanation is that preexisting factors (e.g., low vocabulary, lower educational quality, elevated medication burden, or minority race/ethnicity) not included in the AEG model influenced performance. AEG correction is likely to over-classify MCI in these patients. In contrast, when performances lands in the MCI range for C-model scores but not AEG-model scores, the most likely explanation is the classical crystallized–fluid dissociation: a high-baseline patient (high vocabulary, advanced education, intact crystallized ability) showing below normal performance on tests of fluid abilities.

### 4.4 Race-as-covariate

The C-model included race indicators as covariates in approximately 61% of cognitive measures. However, the boundaries of racial categories are historically and regionally contingent—the 22.3% Other/Mixed race composition of our Bay Area sample underscores that conventional race categories do not partition the population cleanly. Race served in the C-model as a proxy for the constellation of social, educational, and experiential factors associated with racialized identity in the United States (Manly, 2006; Manly & Echemendia, 2007; Brickman, Cabo, & Manly, 2006; Rivera Mindt, Byrd, Saez, & Manly, 2010): differential educational quality not captured by years of education, differential healthcare access, differential occupational opportunity, environmental exposures, chronic stressors, and anxiety during cognitive testing (Steele & Aronson, 1995). Race captures a portion of this variance, and its inclusion in the C-model substantially reduced demographic classification disparities as documented in Section 3.2. The residual Black-vs-White MCI classification ratio of 1.8 was approximately one-third of the AEG ratio (5.6). The C-model difference of approximately four percentage points (9.0% Black vs 5.1% White below the MCI cutoff) likely reflects genuine differences in cognitive risk-factor exposure (vascular disease prevalence, environmental exposures, healthcare access).

The all-or-none rule for race indicators (if any race indicator passed stability selection, all three were retained) avoided selective per-group adjustments that would be difficult to interpret. Under this rule, race-Asian and race-Other factors were retained in 61% of measures but with small coefficients (Asian, mean β = – 0.034, SD 0.179; Other, mean β = – 0.098, SD 0.073) that reached significance primarily in language/lexical and fluency measures, consistent with English being a later acquired language for a some participants in our sample. The pattern is consistent with race indicators capturing measure-specific contextual factors (linguistic background, cultural test-content familiarity) rather than uniform racial corrections.

### 4.5 Limitations

#### The sample is region specific

All participants were recruited in the San Francisco Bay Area. The sample’s high education rate (26.4% post-college) and elevated Asian and Other/Mixed race composition differ from typical US normative samples. The C-model’s coefficients are calibrated on this sample and may not generalize to other regional or international populations. The cross-sample validation reported in Section 3.7 establishes generalizability across cohorts within our sampling region but cannot establish generalizability across regions.

#### External clinical validation is absent

We have not evaluated C-model classifications against gold-standard MCI or dementia diagnoses, in part because such diagnoses are typically made from manual assessments using AEG-corrected normative scores, making the comparison partially circular. Prospective longitudinal studies with biomarker confirmation (CSF, PET amyloid/tau) or clinical-progression endpoints will be needed to evaluate whether C-model classifications predict outcomes more accurately than AEG. The bottom-7-percent classification analyses presented here demonstrate that the choice of correction method substantively changes which subjects are flagged as “at-risk”; whether these classifications translate into improved diagnostic accuracy remains to be established.

#### Advanced disease

The crystallized–fluid dissociation breaks down in syndromes that specifically affect crystallized ability—semantic dementia, primary progressive aphasia, advanced Alzheimer’s disease, and severe depression. In these conditions, vocabulary itself declines and ceases to estimate premorbid ability accurately. This would cause the C-model’s vocabulary covariate to become biased downward, making correction unreliable. This is not unique to the C-model; it is a limitation of all premorbid estimation methods that rely on vocabulary or word-reading performance, including the NART, AMNART, WTAR, and TOPF (Marier et al., 2024). In these populations, collateral information about occupational and educational attainment becomes essential, and measures of premorbid cognitive ability should be interpreted with corresponding caution.

## 5. Conclusion

In comparison with the traditional AEG scoring model, the C-model is distinguished by three features. First, the C-model approximately doubles the variance explained by the model regression, improving the precision of performance estimates. Second, the C-model accounts for racial disparities in performance. Third, the C-model integrates the crystallized–fluid comparison directly into the scoring algorithm, obviating the need for post-hoc score correction and improving the detection of cognitive decline in patients with substantial cognitive reserve. When deployed alongside AEG-corrected scores, agreement between AEG and C-model classifications increases diagnostic confidence, while disagreement carries diagnostic information about over- and under-classification of abnormality. By including the results from both scoring pipelines, the degree of scoring-model concordance becomes an informative signal that can improve the confidence and accuracy of cognitive test evaluation.

## Declarations

### Funding

This work was supported by the National Institute on Aging (NIA) under SBIR grant R44AG097322. The funder had no role in study design, data collection and analysis, decision to publish, or preparation of the manuscript.

### Competing interests

Most authors are employees with Neurobehavioral Systems, Inc. (NBS), the developer of the California Cognitive Assessment Battery (CCAB) described in this manuscript. The authors have no other competing interests to declare.

### Ethics approval and consent

All participants provided written informed consent under protocols approved by the Western Institutional Review Board (WIRB protocol 20201196). All procedures were conducted in accordance with the Declaration of Helsinki.

### Clinical trial registration

This study is registered at https://ClinicalTrials.gov under identifier NCT04800588.

### Data availability

De-identified data underlying these analyses are available from the corresponding author upon reasonable request, subject to data-use agreement consistent with the consent obtained from participants.

### Author contributions

All authors contributed to the conception, design, analysis, drafting, or critical revision of this work, and all authors approved the final version for submission.

## Notes

Supported by NIA R44AG097322.

